# Quantitative Fetal Hemoglobin Measurement: Gazelle™ Point-of-Care Electrophoresis versus Standard HPLC Methodology

**DOI:** 10.64898/2026.09.30.26364074

**Authors:** Thomas N. Williams, Roisin Connon, Rita Muhindo, Denis Amorut, Paul Ongodia, Charles B. Okalebo, Julian Abeso, Florence Alaroker, Sarah Asio, Amos Odiit, Charles Kiyaga, Sophie Uyoga, George Mochamah, Gideon Nyutu, Christabel Mogaka, Asad Muyinda, Grace Abongo, Kathryn Maitland, Diana M Gibb, A. Sarah Walker, Elizabeth George, Russell E. Ware, Peter Olupot-Olupot

## Abstract

Quantitative measurement of fetal hemoglobin (HbF) using high-performance liquid chromatography (HPLC) is standard but expensive and widely unavailable in low-income settings. The newer Gazelle™ method, which employs single-use alkaline electrophoresis cartridges, provides a portable and relatively inexpensive potential alternative. In the current study, we tested 3055 venous blood samples collected from 1780 children with sickle cell anemia who were enrolled into the H-PRIME trial by both Gazelle and the Bio-Rad D-10 HPLC instrument, before and during hydroxyurea treatment. Data reinterpretation was required to quantify accurately high HbF values derived using the D-10. Mean (standard deviation) baseline HbF values by HPLC and Gazelle (N=2042) were 11.4% (SD 5.9%) versus 16.3% (SD 7.5%), respectively, while mean on-treatment HbF values (N=1013) were 18.9% (SD 9.0%) versus 25.4% (SD 11.5%), respectively. While overall agreement was good (Lin’s concordance 0.76, 95% CI 0.75-0.78), HbF values derived using Gazelle were modestly higher than HPLC values (Bland-Altman agreement mean +5.4%, 95%CI −2.4% to +13.2%), with the difference between measures increasing at higher HbF percentages. With strong concordance and more easily interpretable HbF values, Gazelle represents an attractive and inexpensive alternative to HPLC for HbF quantification in the low-income settings where most patients with sickle cell anemia live.

**Key Points:**

1. Point-of-care Gazelle electrophoresis compares favorably to standard HPLC hemoglobin separation to quantify fetal hemoglobin.
2. Gazelle HbF quantitation is accurate, inexpensive, and suitable for use in low-income settings including sub-Saharan Africa.

## INTRODUCTION

Fetal hemoglobin (HbF) is an important modifier of sickle cell anemia as it directly inhibits the polymerization of sickle hemoglobin (HbS) in circulating erythrocytes. Higher endogenous HbF levels are protective against most sickle-related clinical complications, including mortality.^1,2^ Moreover, the induction of HbF is the primary mechanism by which hydroxyurea ameliorates disease severity,^3,4^ and as a consequence several successful gene therapy treatments for sickle cell anemia have been developed that are specifically targeted toward the up-regulation of HbF production.^5,6^

Quantitative methods for the measurement of HbF, typically reported as the percentage of total hemoglobin, or more accurately as (HbF/HbF+HbS), have been available for decades. Commonly used methods include electrophoresis using agarose gels in either acidic or alkaline conditions, which separate the major hemoglobins HbA, HbS, HbC, and HbF, and High-Performance Liquid Chromatography (HPLC) methodologies which allow the separation and quantitation of both major and minor hemoglobins.^7^ Electrophoretic methods are relatively inexpensive (typically around $5-10 per test), while HPLC methods are more costly at around $10-20 USD per test and require equipment that is frequently valued at $30,000-$60,000. Moreover, the interpretation of HPLC results at high concentrations of HbF can be difficult, requiring the reanalysis of software outputs based on expert knowledge. For all these reasons, HPLC is not widely used for the measurement of HbF concentrations in low-income settings. Newer methodologies such as capillary electrophoresis and mass spectrometry have even better resolution but require even more costly equipment and equipment maintenance as well as greater technical training.

The single-use Gazelle™ portable point-of-care electrophoresis platform (Hemex Health Inc, Portland OR)^8^ has the potential to provide HbF quantitation at lower cost ($2-5 per test), without the need for technical interpretation and involves a solid-state and portable base unit that retails at <$1,000. In a recent laboratory-controlled comparison, the Gazelle instrument provided accurate point-of-care HbF quantification at levels relevant to hydroxyurea monitoring,^9^ thereby offering a potential alternative for decentralized laboratory testing. Here, we present data from a large clinical trial in Uganda that shows that the Gazelle method is feasible, accurate and well suited to low-income settings.

## METHODS

This study was conducted with samples collected in the H-PRIME trial (ISRCTN15724013, PACTR202401802272880), a large randomized clinical trial of hydroxyurea and other treatments in ~1800 children with sickle cell anemia aged 1-10 years in Eastern Uganda.^10^ H-PRIME is being conducted at four clinical sites with variable access to laboratory facilities: Mbale Regional Referral Hospital, Atutur District Hospital, Ngora Health Centre IV, and Soroti Regional Referral Hospital.^10^ We measured HbF concentrations by both standard ion-exchange HPLC using the BioRad D-10™ machine (BioRad Laboratories, Hercules, CA, USA) and by Gazelle using venous blood samples collected into EDTA both before and at 3-month intervals after the introduction of hydroxyurea therapy.

The D-10 Short Program reports multiple separation peaks that are labelled by instrument software based on reference points for HbA1c, HbA_2_, and HbF. However, as recognized by the manufacturer in their user manual,^11^ these hemoglobin peaks can be incorrectly interpreted in samples when HbF values exceed 10%. In such samples, the D-10 machine will often report results that are not marked accurately according to the standard hemoglobin variants and must therefore be reinterpreted to yield comprehensible results (see Supplementary Figure 1 for examples). In the first example (Supplementary Figure 1A), the D-10 output has been labelled correctly by the software to yield an easily interpretable result. In the second example, however (Supplementary Figure 1B), the software assigned various generic labels to multiple peaks. Re-analysis of such outputs are required to yield interpretable values. For the purposes of our current analysis, we re-calculated HbF values from the D-10 output tracings based on elution times, re-assigning all peaks eluting between 0.4 and 1.0 minute to the HbF value to capture accurately the entire HbF signal in such samples.

We also measured HbF values in the same samples using the Gazelle instrument, which reports only HbA, HbS, HbC, HbF and HbA_2_ as relative percentages and directly provides a numerical value for each. We excluded fourteen Gazelle values that corresponded to machine errors: 1 in which HbF was recorded as 100% and 13 in which HbF was recorded as 0%. All values from both methods were recorded on case report forms and entered onto the trial database written in REDCap™ (Vanderbilt University, Nashville, Tennessee, USA). HbF percentages from both methods were calculated as HbF/(HbF+HbS) as previously reported.^4^

For statistical analysis, we compared agreement between HbF/(HbF+HbS) measured using the Gazelle and D-10 (recalculated where necessary from the machine output) using Lin’s estimate of concordance, Bland-Altman plots with estimates of mean difference and 95% limits of agreement and reduced major axis regression. Analyses were performed both overall, by site, and for each available timepoint.

## RESULTS AND DISCUSSION

Data from the whole study are summarized in Figure 1. A total of 3055 samples were available for the comparison of HbF values derived using both methods. Samples from all four clinical sites were assayed, ranging from 598 samples from the Ngora site to 1101 from Mbale. Before starting hydroxyurea therapy, 2042 samples had a mean (±SD) HbF value of 11.4% (±5.9%) measured on the D-10 and 16.3% (±7.5%) when measured by Gazelle. During the first 12 months of hydroxyurea treatment, a total of 1013 samples were tested by both methods, with a mean HbF value of 18.9% (±9.0%) and 25.4% (±11.5%), respectively.

**Figure 1.**
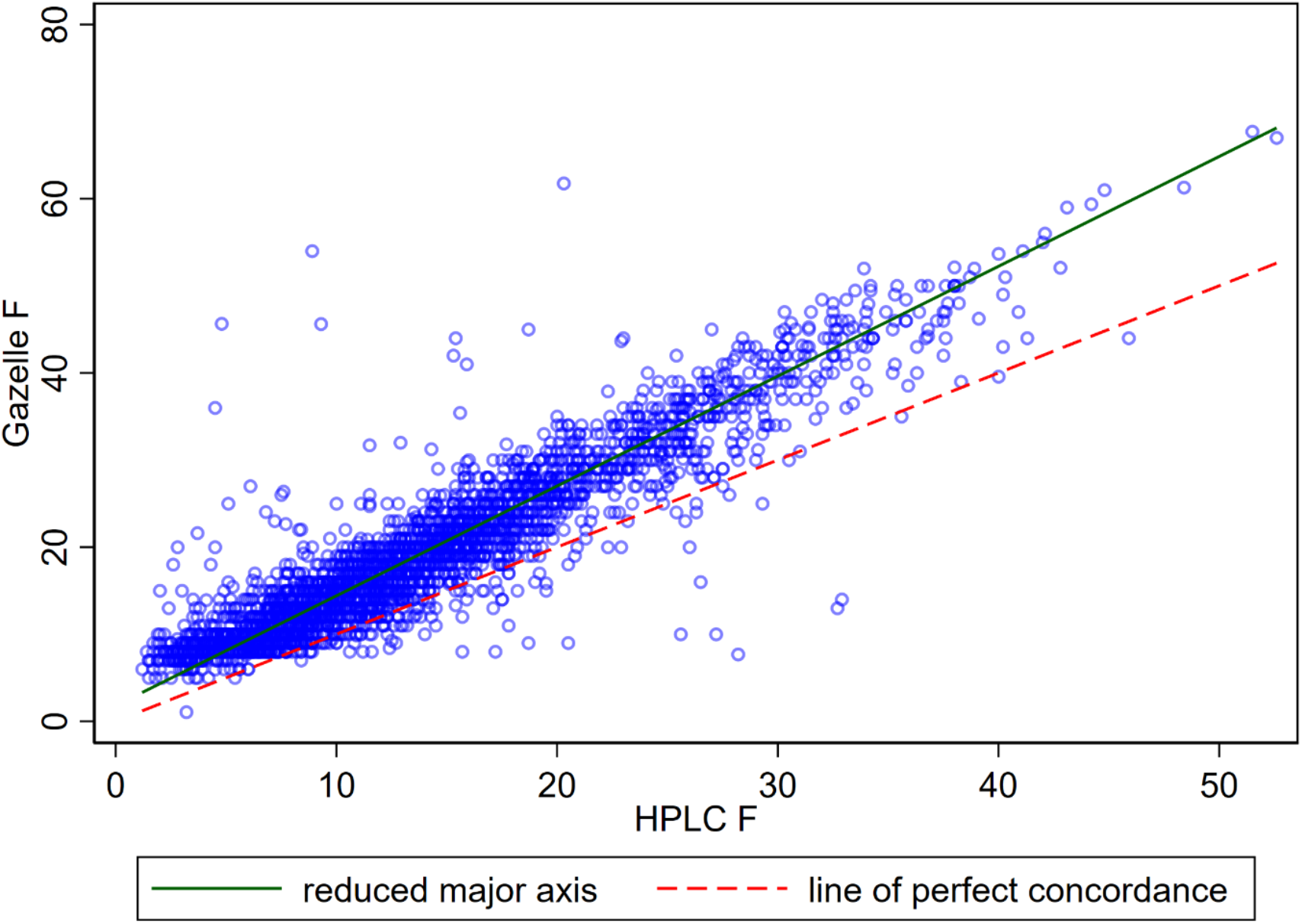
Direct comparison of Gazelle™ HbF versus D-10 HbF values. Scatter plot of Gazelle HbF versus D-10 HPLC HbF values. For the Gazelle we calculated HbF% from the (HbF/HbF+HbS) values that were reported by the machine while for the D-10 we used values recalculated from the machine output as described in the text. The red line shows concordance, if there were perfect agreement between the two methods. The green line is the line of best fit to the observed data. Note: Lin’s concordance coefficient=0.76 (0.75-0.78). Reduced major axis regression shown in red dashed and line of perfect concordance in green solid.

Measurements were well correlated overall, albeit with a best-fit line that differed from unity. The HbF values derived using the Gazelle method were modestly higher than those derived using the D-10, the mean difference being +5.4% and the Bland-Altman 95% limits of agreement being −2.4% to 13.2% (Figure 2). Reduced major axis regression gave an intercept of +1.8% (95% CI 1.6-2.0) and slope of +1.26% (95% CI 1.24-1.28), demonstrating that the difference between the two methods was small at low HbF% values but increased at higher values. Lin’s concordance was 0.76 overall (95% CI 0.75-0.78). The concordance was similar across the 4 sites (ranging from 0.72-0.81), and at each time point (0.68-0.75), as were the overall mean differences (Table 1).

**Table 1.** Mean difference and Lin’s concordance.

|  | Mean difference<br>(Gazelle-HPLC) (%) | 95% limits of agreement | Lin's concordance<br>coefficient (95% CI) |
| --- | --- | --- | --- |
| **Overall** | 5.4 | -2.4, 13.2 | 0.76 (0.75-0.78) |
| **By site** |  |  |  |
| Mbale | 5.5 | -1.9, 12.9 | 0.75 (0.73-0.77) |
| Soroti | 5.8 | -1.3, 12.9 | 0.78 (0.76-0.80) |
| Atutur | 6.5 | -1.7, 14.7 | 0.72 (0.70-0.75) |
| Ngora | 3.7 | -4.2, 11.6 | 0.81 (0.79-0.83) |
| **By timepoint** |  |  |  |
| Screening | 4.7 | -1.9, 11.4 | 0.69 (0.68-0.71) |
| Month 0 | 5.8 | -0.5, 12.2 | 0.68 (0.64-0.73) |
| Month 3 | 6.8 | -2.4, 16.0 | 0.72 (0.69-0.76) |
| Month 6 | 6.3 | -3.3, 15.9 | 0.75 (0.71-0.78) |
| Month 12 | 6.6 | -2.7, 15.9 | 0.75 (0.72-0.79) |

**Figure 2.**
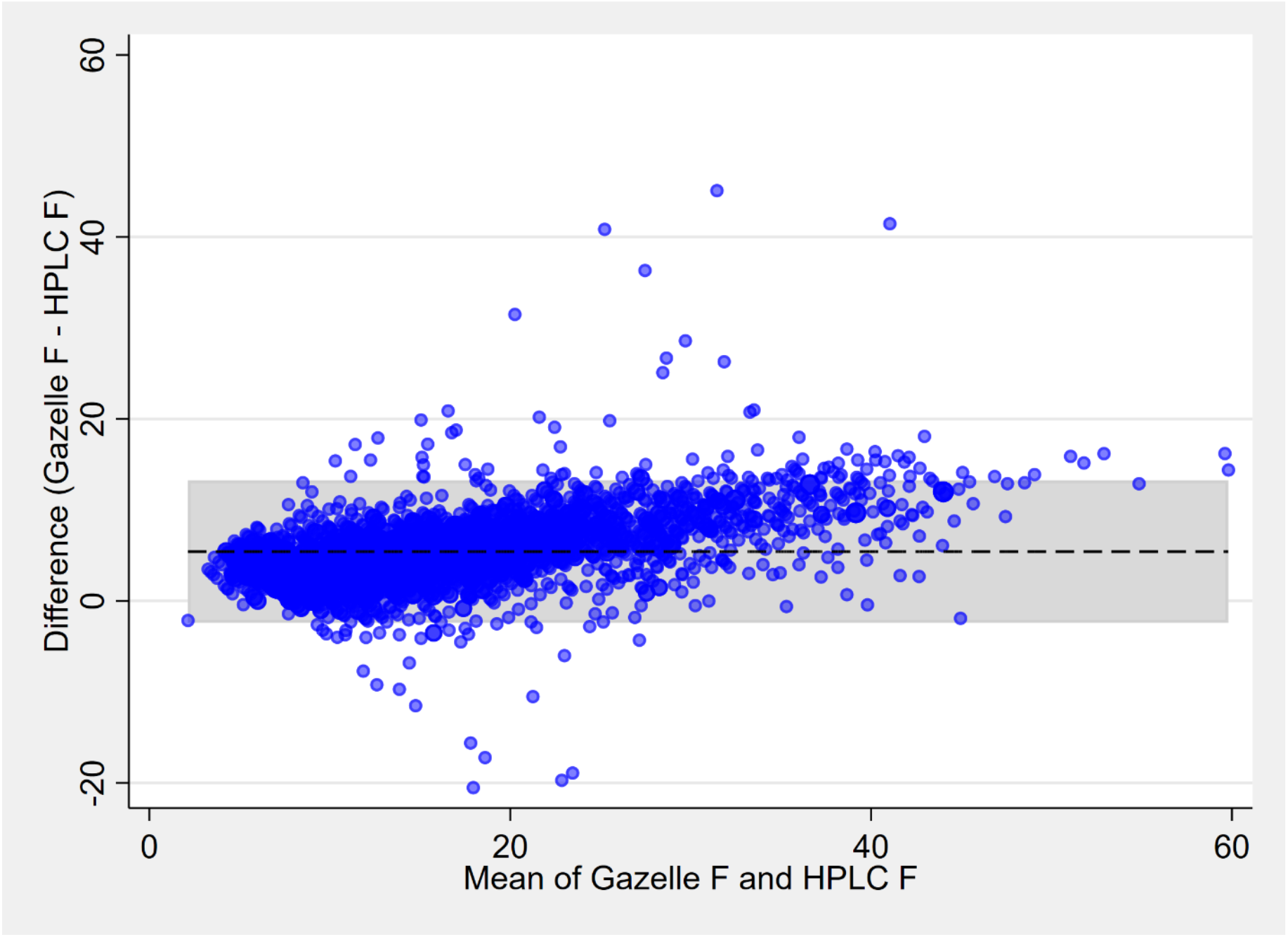
Bland-Altman plot showing the difference in HbF% values derived using the Gazelle and D-10 methods. Bland-Altman plot showing the difference between the Gazelle-HPLC HbF values versus the average of the two methods. The dashed line is the mean difference between the measures, and the shaded area is the range containing 95% of the differences. Dashed line shows the mean difference between the measures, and the shaded area is the range in which 95% of the differences are predicted to fall)

Induction of HbF is an important mechanism for hydroxyurea therapy in children and adults with sickle cell anemia, and monitoring HbF trends is therefore often used assess both the treatment response and compliance with medication. HbF quantitation is typically performed in centralized laboratories using expensive equipment and highly trained personnel, which can be challenging in the low-income settings in sub-Saharan Africa where the burden of sickle cell anemia is highest.^12^ The Gazelle instrument provides single-use hemoglobin electrophoresis using a portable point-of-care platform, and was recently shown to yield similar HbF% results to both HPLC and capillary zone electrophoresis when used in a controlled laboratory environment.^9^ However, before this study, this system had not been tested and formally compared to HPLC in a low-income setting.

Gazelle showed good overall concordance of HbF% with D-10 values (Figure 1), although Gazelle produced slightly higher values with increasing HbF concentrations (Figure 2). Although the D-10 instrument is often considered the “gold standard” for HbF quantification, it may not be accurate at elevated values. As noted above, the Short Program package insert notes difficulties with reporting accurate HbF measurements in samples with an HbF value of >10%,^11^ but as shown in Supplementary figure 1, reinterpretation of the D-10 tracings can more accurately quantify the HbF value. Such data manipulation would not be practical in busy and understaffed hospitals in many parts of sub-Saharan

Africa. By contrast, the Gazelle outputs data that is much more easily interpreted and which does not require re-calculation. After sample loading, both assays are completed and ready for interpretation in 8-10 minutes.

The need for portable and inexpensive point-of-care methods for the measurement of HbF in patients with sickle cell anemia is well-recognized.^13,14^ Given the strong concordance, rapid results and cheaper cost, Gazelle is an attractive option for monitoring HbF concentrations in patients with sickle cell anemia who are being treated with hydroxyurea in Africa. With recent evidence supporting the safety and efficacy of long-term hydroxyurea treatment of sickle cell anemia,^15,16^ simple and cost-effective methods for monitoring the HbF response to hydroxyurea therapy will become increasingly important.

Quantitative HbF measurement using Gazelle offers a simple, robust, reliable, affordable, and accurate alternative to standard HPLC testing.

## Data Availability

All data produced in the present study are available upon reasonable request to the authors

## ACKNOWLEDGMENTS

We thank the children and families who are participating in the trial and the clinical, nursing, fieldworker, pharmacy and laboratory teams that have supported this study across the four sites including George Masifa, Peter Shisisa, Emma Khisa, Edrin Mwanda, Faizo Machoka, Joyce Khwaka, Dominic Wabwire, Timothy Derrick Musansala, Hope Idionyi, Stanley Ecuru, Benjamin Otim, Heather Egau, Erasmus Otim and Juventine Oriada. We thank Hemex Health for providing the equipment and reagents used in confirmatory testing for sickle cell anemia and the measurement of HbF concentrations.

## AUTHOR CONTRIBUTIONS

Study design: TNW, RC, CM, ASW, EG, REW, P-OO

Study conduct: RM, DA, PO, CBO, JA, FA, SA, AO, CK, SU, GM, GN, AM, P-OO

Data interpretation and analysis: TNW RC, RM, GM, GN, CM, GA, ASW, EG, REW, P-OO Write-up: all authors

Funding: TNW, JA, FA, SA, SU, KM, DMG, ASW, EG, REW P-OO

---

**Supplementary Figure 1.**
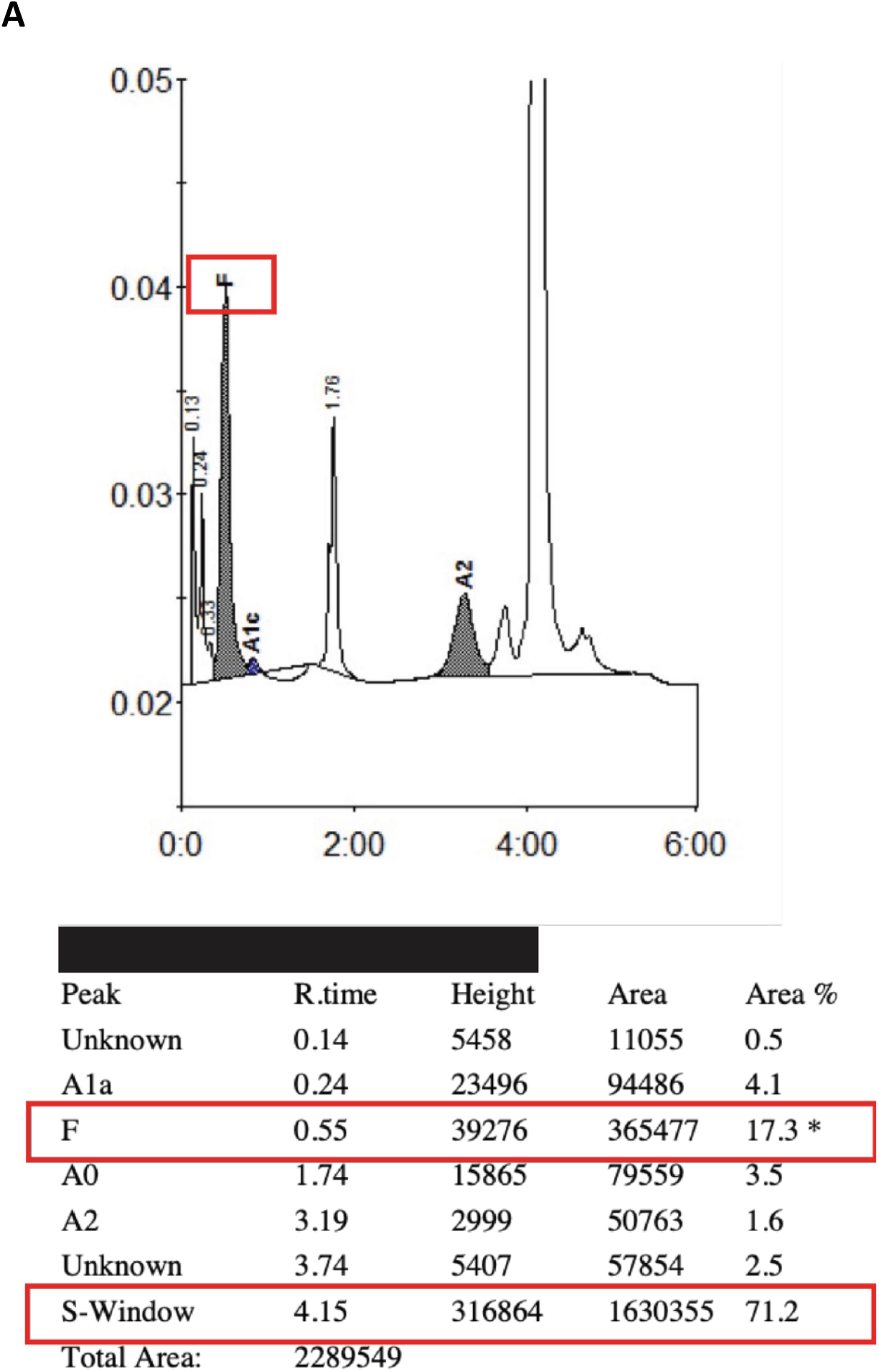
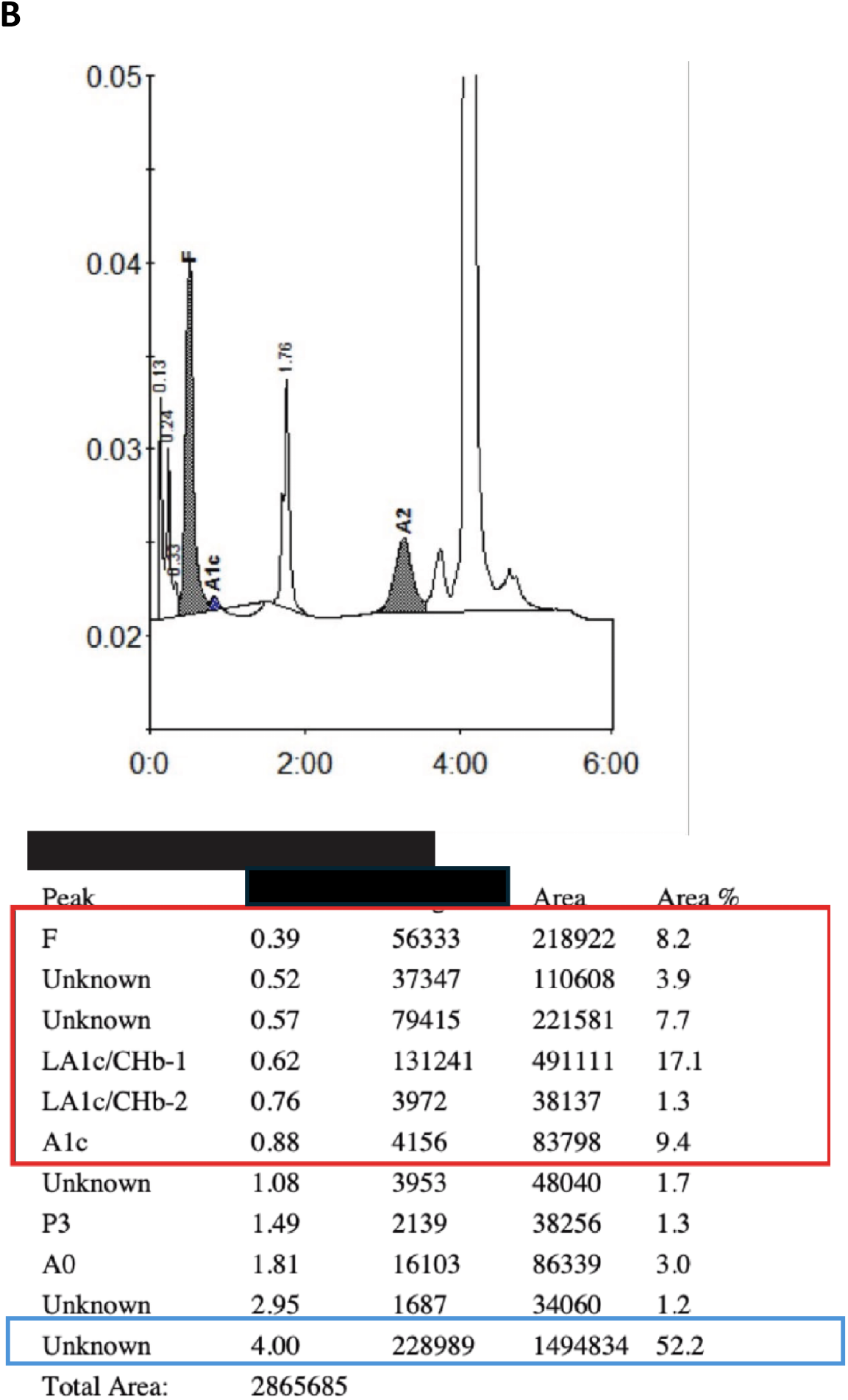
Example outputs from the D-10 machine. In this case, the output from the D-10 software is easily interpreted. The software labelled the peaks appropriately so that result reported for HbF include all peaks eluting between 0.4-1.0 minute while the HbS peak captures all material eluting around 4 minutes. HbF% was calculated as HbF/HbF+HbS using the areas under the corresponding curves. In this case, the total area under the HbF curves was 365477 and the total area under the HbS curve was 1630355. The total area under the two curves combined was therefore 2658991, yielding an HbF value of 18.3% and HbS value of 81.7%. In this case, the D-10 software did not label the peaks appropriately. Six peaks are listed between 0.4 and 1.0 minute, only one of which is labeled HbF and the large HbS peak at 4 minutes is labeled as unknown. In such cases, for the purpose of analysis we re-calculated HbF values by summing the areas under all peaks eluting between 0.4 and 1 minute (column labelled R time) and HbS values by summing all areas under peaks eluting after 4 minutes. All other areas were disregarded. The HbF% was then reported using the recalculated values for HbF/HbF+HbS. In this case, the total area under the HbF curves (red box) was 1164157 and the total area under the HbS curve (blue box) was 1494834. The total area under the two curves combined was therefore 2658991, yielding an HbF value of 43.8% and an HbS value of 56.2%.

